# Determinants associated with infant mortality in Ethiopia: Using the recent 2019 Ethiopia mini demographic and health survey

**DOI:** 10.1101/2023.02.14.23285933

**Authors:** Yeshambel Kindu Yihuna, Abay Kassie Lakew, Nigist Mulu Takele, Seyoum Woldebrehan Agelu, Adane Agegn Enigda

## Abstract

**Background:** Infant mortality is the number of deaths under the age of one year and it is one of the most essential and sensitive indicators of the health status of the country. This study aims to identify the determinants that affect infant deaths in Ethiopia using the 2019 EMDHS.

**Methods:** This study used the 2019 Ethiopia mini demographic and health survey and 1,397 infants born from mothers who had been interviewed about births in the five years before the survey. The data were first analyzed with a chi-square test of association, and then potential factors were evaluated with binary logistic regression models. An adjusted odds ratio (AOR) along with a 95 % confidence interval (CI) of parameters was used to interpret the result

**Results:** The prevalence of infant death was 21%. The study also showed that age at first birth (16-32) infant (AOR = 0.541, 95%CI: 0.353, 0.827), mothers who had breastfed (AOR = 0.022, 95% CI: 0.014,), preceding birth interval less than 24 months (AOR = 0.183, 95% CI 0.117, 0.287), mothers who born their child in the health sector (AOR = AOR = 0.442, 95% CI: 0.304, 0.802) statistically related with a lower risk of infant mortality. Although mothers age group 35-49 (AOR = 2.682 1.446 4.974), mothers who had no ANC visits (AOR = 5.029, 95% CI: 2.923, 8.64), multiple births (AOR = 6.094, 95% CI: 2.684, 13.837, P=0.000) were statistically associated with a higher risk of infant mortality.

**Conclusions:** It is greatly suggested that maternal and child health care services (ANC visits) are strengthened. Preceding birth interval of fewer than 24 months, multiple births, and please of delivery at home needs special attention. We recommend also health institutions play a great roll to give awareness to mothers about family planning to reduce infant mortality.

## Introduction

Mortality is very responsive to social, economic, and psychological factors. Historically, mortality has often been used as a barometer of welfare **[1]**. Under-five mortality is the most sensitive indicator of the health status of the community in the world. Despite their having been a significant reduction in under-five mortality, its rate is still high in Sub-Saharan African countries [**2**]. Infant mortality is the number of deaths under the age of one year and it is one of the most essential and sensitive indicators of the health status of the country **[3]**. Infant mortality is a useful and inexpensive indicator of population health. It represents not only the health of newborns but also the general well-being of society**[4]**. Neonatal time is the highest risk for infant mortality because of pre-term birth (28 %), severe infections (26%), and asphyxia (23%) **[5]**. In Ethiopia, infant and child mortality is still high. According to the 2019 EMDHS report in Ethiopia 1 in every 30 children dies within the first month, 1 in every 22 children dies before celebrating their first birthday, and 1 in every 17 children dies before reaching their fifth birthday. The report also showed that infant mortality also declined from 59 deaths per1, 000 live births in 2011 to 48 deaths per1, 000 live births in 2016 and declined from 48 deaths per1, 000 live births in 2016 to 47 deaths per 1,000 live births in 2019 [**6-8**]. There has been a slight increase in neonatal mortality since 2016, from 29 to 33 deaths per 1,000 live births **[8]**. Infant mortality was still high in Africa including Ethiopia and it can be reduced by increasing the mother’s age at first birth, duration of breastfeeding, and birth interval **[9]**.The study carried out in different parts of the world **[10-12]** revealed that the combined effect of Maternal age, maternal age at birth, breastfeeding status, type of birth, preceding birth interval, the breastfeeding status, ANC visits, type of birth, and place of delivery, were significant factor for infant death. These studies contributed to our understanding of many features of determinants of infant and child mortality in the world especially, in Ethiopia. This study studied in different ways, child and mother’s demographic and socio-economic variables. This study aimed to identify determinants associated with infant mortality in Ethiopia. However, In Ethiopia as far as our knowledge there is no study done on infant mortality using EMDHS2019 by application of the binary logistic regression model. Moreover, the previous study in Ethiopia was limited to regional survey data. Therefore, this study focuses on identifying the determinants related to infant mortality in Ethiopia using the EMDHS 2019.

## Material and Methods

### Study area and data source

The data used in this study was obtained from the 2019 Ethiopia Mini Demographic and Health Survey (EMDHS 2019), which was implemented by the Central Statistical Agency (CSA) at the request of the Federal Ministry of Health (FMoH). This community-based cross-sectional study was carried out from March 21, 2019, to June 28, 2019, based on a nationally representative sample that provided estimates at the national and regional levels and for urban and rural areas. The survey interviewed 8,855 women of reproductive age (age 15-49) from a nationally representative sample of 8,663 households. This survey was conducted at the national level and for Ethiopia’s nine regional states and two city administrations.The data used for infant mortality estimation were collected in the birth history section of the woman’s questionnaire which is included in the survey. A detailed methodology has been presented in the 2019 EMDHS final report **[13]**.

### Inclusion and exclusion criteria

In this study, a child with an age of less than one year was included. Children older than 1 year are not included in this study. Moreover, our analysis excludes infants whose events (alive or dead) were missing.

### Variables in the study

Outcome variable of this study was infant death in months. Infant mortality is the number of deaths under the age of one. The response variable is coded as (1=alive and 0=death)

Independent variables. The Predictable independent variables that were considered in this study were shown (**Table 1**).

**Table 1.**
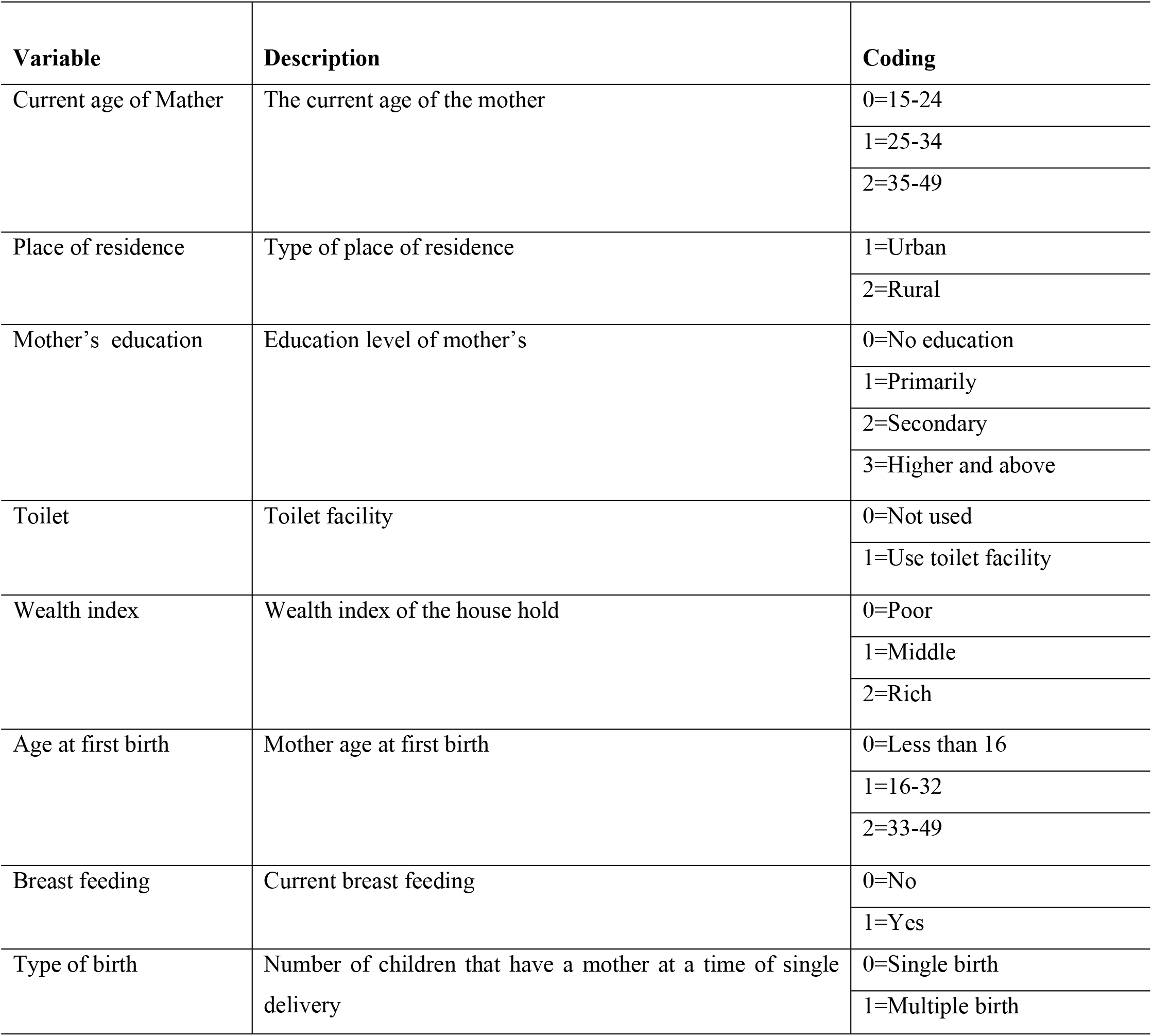
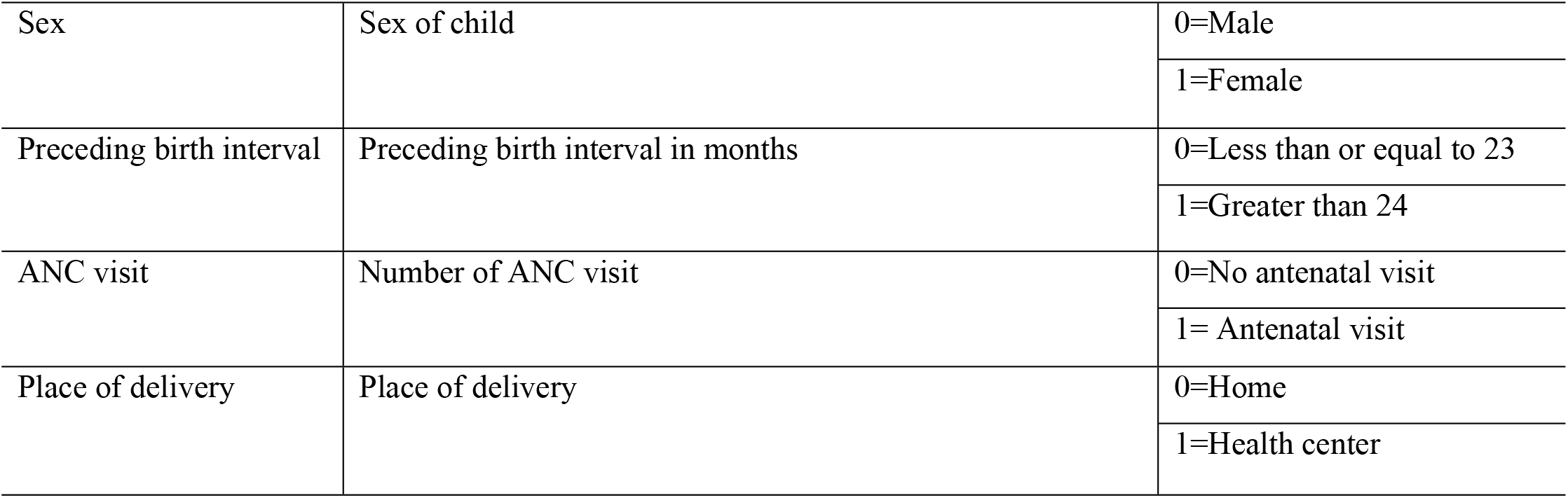
Describition of predictors and coding

We extracted the dependent variable infant mortality rate by using SPSS version 23. After extracting data, Statistical analysis was performed using STAT version 14. To describe the study participants, descriptive statistics were used. This study also used a combination of the chi-square test to determine whether the response variable was associated with different independent variables. Additionally, the multivariable binary logistic regression model performs a response variable with two categories (infant mortality with two categories of yes or no). A binary logistic regression model was used to determine the factors related to infant mortality. A p-value < 5% is used to test the presence of a significant association between the response variable and predictors.

## Result

A total of 1,397 infant mortality took part in this study and infant death occurred in 21 %(293). Rural residents had the highest infant mortality (16.5%).Moreover most women (39.7%) are economically poor. Mother’s age at first birth between 33 and 49 had the lowest rate of infant death (0.8%). The proportion of female and male infant death was (39.4% and 39.6%) respectively. The majority of (38.7%) infant death was attributed to women who didn’t have education Furthermore; all covariates are presented in (**Table 2**).

**Table 2.**
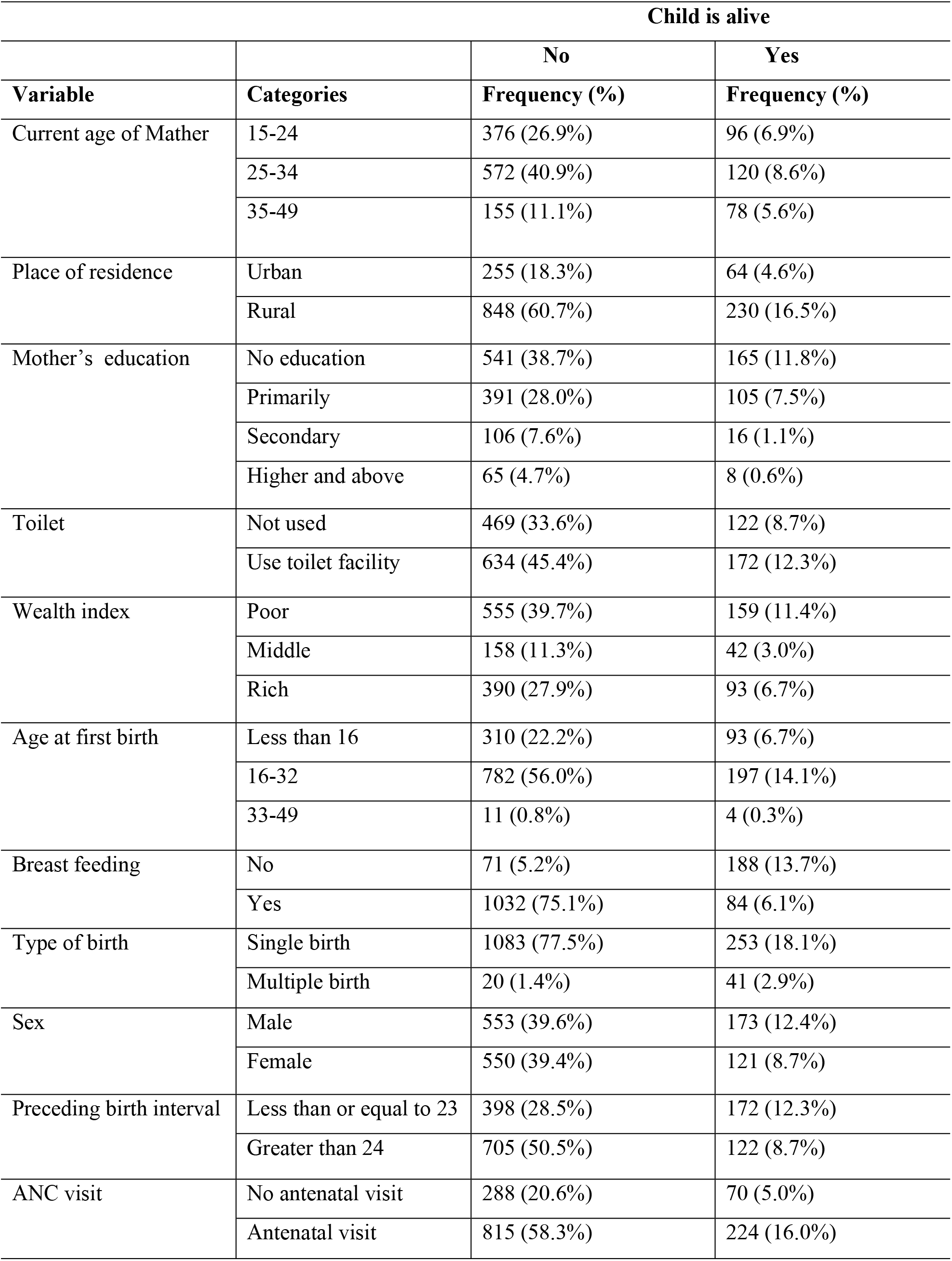
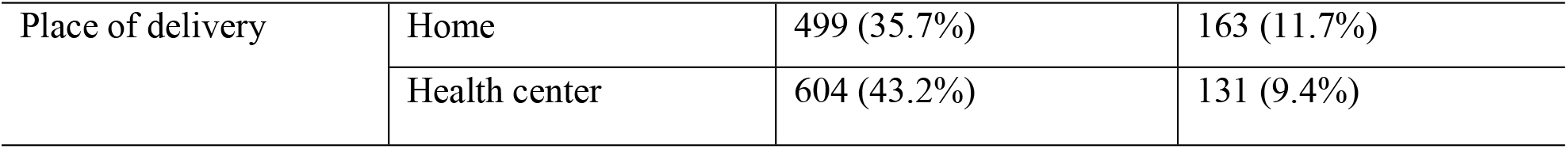
Summary results of independent variables of infant mortality (EMDHS 2019)

| **Child is alive** |  |  |  |
| --- | --- | --- | --- |
| **Variable** | **Categories** | **No**<br>**Frequency (%)** | **Yes**<br>**Frequency (%)** |
| Current age of Mather | 15-24 | 376 (26.9%) | 96 (6.9%) |
|  | 25-34 | 572 (40.9%) | 120 (8.6%) |
|  | 35-49 | 155 (11.1%) | 78 (5.6%) |
| Place of residence | Urban | 255 (18.3%) | 64 (4.6%) |
|  | Rural | 848 (60.7%) | 230 (16.5%) |
| Mother's education | No education | 541 (38.7%) | 165 (11.8%) |
|  | Primarily | 391 (28.0%) | 105 (7.5%) |
|  | Secondary | 106 (7.6%) | 16 (1.1%) |
|  | Higher and above | 65 (4.7%) | 8 (0.6%) |
| Toilet | Not used | 469 (33.6%) | 122 (8.7%) |
|  | Use toilet facility | 634 (45.4%) | 172 (12.3%) |
| Wealth index | Poor | 555 (39.7%) | 159 (11.4%) |
|  | Middle | 158 (11.3%) | 42 (3.0%) |
|  | Rich | 390 (27.9%) | 93 (6.7%) |
| Age at first birth | Less than 16 | 310 (22.2%) | 93 (6.7%) |
|  | 16-32 | 782 (56.0%) | 197 (14.1%) |
|  | 33-49 | 11 (0.8%) | 4 (0.3%) |
| Breast feeding | No | 71 (5.2%) | 188 (13.7%) |
|  | Yes | 1032 (75.1%) | 84 (6.1%) |
| Type of birth | Single birth | 1083 (77.5%) | 253 (18.1%) |
|  | Multiple birth | 20 (1.4%) | 41 (2.9%) |
| Sex | Male | 553 (39.6%) | 173 (12.4%) |
|  | Female | 550 (39.4%) | 121 (8.7%) |
| Preceding birth interval | Less than or equal to 23 | 398 (28.5%) | 172 (12.3%) |
|  | Greater than 24 | 705 (50.5%) | 122 (8.7%) |
| ANC visit | No antenatal visit | 288 (20.6%) | 70 (5.0%) |
|  | Antenatal visit | 815 (58.3%) | 224 (16.0%) |
| Place of delivery | Home | 499 (35.7%) | 163 (11.7%) |
|  | Health center | 604 (43.2%) | 131 (9.4%) |

### Results of the binary logistic regression model

Binary logistic regression model were performed utilization of categorical independent variables that were found to be significant in the bivariate analysis, **Table 4** summarizes the findings. Maternal age, maternal age at birth, breastfeeding status, type of birth, preceding birth interval, breastfeeding status, ANC visits, type of birth, and place of delivery, were statistically significant related with infant death. When compared to maternal age at first birth infants whose age at first birth were (16-32) than whose age less than 16 to infant age, the odds of infant death were 0.541 times (AOR = 0.541, 95%CI: 0.353, 0.827) lower. Those who had breastfed a 0.022 (AOR = 0.022, 95% CI: 0.014, 0.034, P<0.000) times less risk of infant death compared to other infants. The odds of infant death among multiple births were 6.094 (AOR = 6.094, 95% CI: 2.684, 13.837, P=0.000) times higher as compared to single births. Furthermore, Children born greater than 24 months of the previous birth interval had 0.183 times (AOR = 0.183, 95% CI 0.117, 0.287) less risk of infant death than those born less than 24 months.

Compared to mothers who had ANC visits, the risk of infant mortality was 5.029 times higher (AOR = 5.029, 95% CI: 2.923, 8.649, P=0.000). Please of delivery was also a significant predictor of Infant death. Mothers who birth their child in the health sector had a 0.494 (AOR = AOR = 0.442, 95% CI: 0.304, 0.802) times lower risk of infant death than mothers who birth their children at home (**Table 3**).

**Table 3.** Logistic regression model for infant death in Ethiopia (EMDHS2019)

| Variables | B | S.E | Sig. | Exp (B) | 95% CI for Exp (B) |  |
| --- | --- | --- | --- | --- | --- | --- |
|  |  |  |  |  | Lower | Upper |
| Current age of mother (25-34) | 0.238 | 0.305 | 0.321 | 1.269 | 0.792 | 2.032 |
| Current age of mother (35-49) | 0.986 | 0.845 | 0.002 | 2.682 | 1.446 | 4.974 |
| Place of residence (Rural) | 0.074 | 0.311 | 0.796 | 1.077 | 0.611 | 1.897 |
| Mother's education (Primary) | -0.248 | 0.182 | 0.287 | 0.779 | 0.492 | 1.233 |
| Mother's education(Secondary) | -0.697 | 0.218 | 0.112 | 0.497 | 0.211 | 1.176 |
| Mother's education (Higher) | -0.809 | 0.241 | 0.135 | 0.445 | 0.153 | 1.287 |
| Toilet (Use toile) | 0.053 | 0.245 | 0.819 | 1.054 | 0.668 | 1.663 |
| Wealth index (Middle) | 0.203 | 0.381 | 0.513 | 1.225 | 0.666 | 2.252 |
| Wealth index (Rich) | 0.298 | 0.402 | 0.316 | 1.348 | 0.751 | 2.418 |
| Age at first birth (16-32) | -0.614 | 0.117 | 0.005 | 0.541 | 0.353 | 0.827 |
| Age at first birth (33-49) | -1.428 | 0.223 | 0.126 | 0.239 | 0.038 | 1.491 |
| Breast feeding (Yes) | -3.801 | 0.005 | 0.000 | 0.022 | 0.014 | 0.034 |
| Type of birth (Multiple) | 1.807 | 2.549 | 0.000 | 6.094 | 2.684 | 13.837 |
| Sex of child (Female) | -0.284 | 0.144 | 0.138 | 0.752 | 0.516 | 1.095 |
| Preceding birth interval (>24) | -1.693 | 0.042 | 0.000 | 0.183 | 0.117 | 0.287 |
| ANC visit (Antenatal.) | 1.615 | 1.391 | 0.000 | 5.029 | 2.923 | 8.649 |
| Place of delivery(health sector) | -0.703 | 0.122 | 0.004 | 0.494 | 0.304 | 0.802 |

### Assessment of goodness of fit of the model

After fitting the binary logistic regression model to categorical data, it is necessary to assess the adequacy of the model. To deal with this, we used Hosmer and Lemeshow’s Goodness of Fit tests. Based on the results in (Table 4), the null hypothesis that there is no difference between the model with just a constant and the model with predictors has been rejected. The statistical values **(Table 5)** of Cox & Snell (Pseudo R-square = 0.529) and Nagelkerke (Pseudo R-square = 0.839) were reasonable in this investigation, indicating that the model explained part of the variation **(Table 6). Table 7** reflects that the Hosmer-Lemeshow goodness of fit test is an insignificant result. As a result, the model well fits the data.

**Table 5.** Omnibus tests of model coefficients.

|  | Chi-square | Df | Sig. |
| --- | --- | --- | --- |
| Step | 1035.037 | 14 | 0.000 |
| Block | 1035.037 | 14 | 0.000 |
| Model | 1035.037 | 14 | 0.000 |

**Table 6.** Model Summary.

| Step | 2Loglikelihood | Cox & Snell R Square | Nagelkerke R Square |
| --- | --- | --- | --- |
| 1 | 332.711<sup>a</sup> | 0.529 | 0.839 |

**Table 7.** Hosmer and Lemeshow test

| Chi-square | Df | Sig. |
| --- | --- | --- |
| 4.439 | 8 | 0.82 |

## Discussion

This study aims to identify the determinants of infant deaths using EMDHS 2019. To examine factors that affect infant mortality, this study used both descriptive and logistic regression models. The descriptive statistics showed that the prevalence of infant death was 21%. The binary logistic regression model shows that maternal age, age of mothers at birth, breastfeeding status, type of birth, preceding birth interval, breastfeeding status, ANC visits, type of birth, and place of delivery, were statistically significantly related to infant deaths. Mother’s age at first birth is a significant predictor of infant death. Infant death was higher in mother’s birth their child at young stage than in the older mother. This outcome is consistent with study **[9,14-16]**. This finding was consistent with a study from the year before **[28]**. This study finding has important policy implications, particularly in determining the program needs for a sustainable decline in infant mortality rate, and in monitoring public health interventions. The mother’s age was found to be a significant predictor of infant death. Infant death was higher in the younger mother than in the older mother. This study is consistent with findings **[17-18]**. The findings also revealed that type of birth was a significant risk factor for infant mortality. The risk of infant death was greater in multiple births than in single birth infant. This is because single newborns get enough breast milk **[19-21]**. These studies **[17,22]**. also agree with the previous study [Breastfeeding is a child’s first line of defense against death and disease, protecting them from respiratory infections, gastrointestinal illnesses, and other negative health consequences **[9]**. Short birth intervals had high risk of infant death than long birth intervals, and the risk of infant death decreased as the prior birth interval grew. The risk of obstetric complications is lower in mothers with long birth intervals than in mothers with shorter birth intervals **[21-23]**. Furthermore, statistically, place of delivery was also a significant predictor of infant death. Mothers who birth their child in the health sector had a lower risk of infant death than mothers who birth their children at home.This is because newborns health sector have greater access to health care and other key health-related amenities that are required for infant survival **[24-27]**. It is important to look beyond identifying determinates with infant death. Reducing motherhood in younger ages and increasing the spacing between births and also specified that an increase in mothers’ age at first birth, improve health care services which should in turn raise child survival and should decrease infant death in Ethiopia. Interventions and strategies should be targeted focusing on these characteristics to improve child health outcomes as well as the future betterment of Ethiopia.

## Conclusion

The result of this study revealed that maternal age, mothers’ age at birth, breastfeeding, type of birth, preceding birth interval, breastfeeding status, ANC visits, type of birth, and place of delivery, were statistically significantly related to infant mortality. Preceding birth interval of fewer than 24 months, multiple births, and please delivery at home needs special care. We recommend also health institutions play a great roll to give awareness to mothers about family planning and breastfeeding to reduce infant mortality.

## Data Availability

Availability of data .All relevant data are within the paper and its. Supporting information files

http://www.dhsprogram.com.

## Acknowledgments

The authors would like to thank the measure DHS program for permitting access to the EMDHS data sets.

## Author Contributions

YK creation the data, formal analysis, writing original draft and supervision, AK and NM make validation, SW visualization, AA and YK writing the review and all authors review the final manuscript.

## References

1. Singatiya, M. (2013). “Socioeconomic determinants of infant mortality rate in India.” The Journal of Public Health: Photon 115: 163–174.

2. Yalew, M., et al. (2022). “Time to under-five mortality and its predictors in rural Ethiopia: Cox-gamma shared frailty model.” Plos one 17(4): e0266595

3. Hunter, B. M. and J. D. Shaffer (2022). “Human capital, risk and the World Bank’s reintermediation in global development.” Third World Quarterly 43(1): 35–54.

4. Reidpath, D. D. and P. Allotey (2003). “Infant mortality rate as an indicator of population health.” Journal of Epidemiology & Community Health 57(5): 344–346.

5. Masuy-Stroobant, G. 2002. The determinants of infant mortality: how far are conceptual.

6. Demographic E. Health survey 2011 central statistical agency Addis Ababa. Maryland, USA: Ethiopia ICF International Calverton; 2012.70–71p.

7. Demographic E. Health Survey Central Statistical Agency and ICF International Addis Ababa. Maryland, USA: Ethiopia ICF International Calverton; 2016.

8. Demographic E. Health Survey Central Statistical Agency and ICF International Addis Ababa. Maryland, USA: Ethiopia ICF International Calverton; 2019

9. Mulugeta, S. and S. Wassihun (2022). “Determinant of infant mortality in Ethiopia: demographic, socio economic, maternal and environmental factors.” MOJ Women’s Health 11(2): 49–57.

10. Kotsadam A., et al., Development aid and infant mortality. Micro-level evidence from Nigeria. World Development, 2018. 105: p. 59–69.

11. Geruso, M. and D. Spears (2018). Heat, humidity, and infant mortality in the developing world, National Bureau of Economic Research.

12. Gonzalez, R. M. and D. Gilleskie (2017). “Infant mortality rate as a measure of a country’s health: a robust method to improve reliability and comparability.” Demography 54(2): 701–720.

13. Ephi, I. (2019). “Ethiopian Public Health Institute (EPHI)[Ethiopia] and ICF.” Ethiopia Mini Demographic and Health Survey 2019: Key Indicators.

14. Khan, J. R. and N. Awan (2017). “A comprehensive analysis on child mortality and its determinants in Bangladesh using frailty models.” Archives of Public Health 75(1): 1–10.

15. Kumar, P. P. and G. File (2010). “Infant and child mortality in Ethiopia: a statistical analysis approach.” Ethiopian Journal of Education and Sciences 5(2).

16. Zewdie, S. A. and V. Adjiwanou (2018). “Multilevel analysis of infant mortality and its risk factors in South Africa.” International Journal of Population Studies 3(2): 43–56.

17. Muluye, S. and E. Wencheko (2012). “Determinants of infant mortality in Ethiopia: A study based on the 2005 EDHS data.” Ethiopian Journal of Health Development 26(2): 72–77.

18. Tura, G. (2009). “Antenatal care service utilization and associatedfactors in Metekel Zone, Northwest Ethiopia.” Ethiopian Journal of Health Sciences 19(2).

19. Jarde, A., et al. (2021). “Risk factors of infant mortality in rural The Gambia: a retrospective cohort study.” BMJ paediatrics open 5(1).

20. Fenta, S. M. and H. M. Fenta (2020). “Risk factors of child mortality in Ethiopia: application of multilevel two-part model.” Plos one 15(8): e0237640.

21. Kiross, G. T., et al. (2021). “Individual-, household-and community-level determinants of infant mortality in Ethiopia.” Plos one 16(3): e0248501.

22. Bhusal, M. K. and S. P. Khanal (2022). “A Systematic Review of Factors Associated with Under-Five Child Mortality.” BioMed Research International 2022.

23. Wegbom, A. I., et al. (2019). “Survival analysis of under-five mortality and its associated determinants in Nigeria: evidence from a survey data.” International Journal of Statistics and Applications 9(2): 59–66.

24. Talukder, A., et al. (2021). “Prevalence and Factors Associated with under-5 Mortality in Nigeria: Evidence from 2018 Nigeria Demographic and Health Survey.” Dr. Sulaiman Al Habib Medical Journal 3(4): 154–161.

25. Flannery, D. D., et al. (2021). “Assessment of maternal and neonatal cord blood SARS-CoV-2 antibodies and placental transfer ratios.” JAMA pediatrics 175(6): 594–600.

26. Mulugeta, S. S., et al. (2022). “Multilevel log linear model to estimate the risk factors associated with infant mortality in Ethiopia: further analysis of 2016 EDHS.” BMC Pregnancy and Childbirth 22(1): 1–11.

27. Weldearegawi, B., et al. (2015). “Infant mortality and causes of infant deaths in rural Ethiopia: a population-based cohort of 3684 births.” BMC public health 15(1): 1–7.

28. Fagbamigbe, A., et al. (2021). “Approximation of the Cox survival regression model by MCMC Bayesian Hierarchical Poisson modelling of factors associated with childhood mortality in Nigeria.” Scientific Reports 11(1): 1–18.

